# Electrocardiographic abnormalities and Cardiovascular outcomes in systemic autoimmune rheumatic diseases: A scoping review protocol

**DOI:** 10.1101/2025.07.06.25330954

**Authors:** Adriana Munford Lima Pimentel, Alfredo dos Santos Ribeiro, Anamaria Siqueira Han, Isabela Silva Erthal Vieira, Leonardo Cardoso dos Santos, Lucas Chein Ferreira, Manuela Luz Loureiro, Silvia Marina de Amorim Figueira

**Affiliations:** Universidade Federal Fluminense, Rio de Janeiro, RJ, Brazil; Institute D’Or for Research and Education (IDOR), Rio de Janeiro, RJ, Brazil

**Keywords:** “electrocardiogram”, “rheumatic diseases”, “autoimmune diseases”

## Abstract

**Objective:** This Scoping review seeks to carry out a global mapping of the literature on electrocardiographic abnormalities and major adverse cardiovascular outcomes in patients with systemic autoimmune rheumatic diseases.

**Introduction:** Systemic autoimmune rheumatic diseases (SARD) constitute a heterogeneous group of inflammatory disorders. Cardiovascular involvement in SARD may result from the involvement of different structures, including the electrical system, leading to rhythm and conduction disturbances. Identifying electrocardiographic changes associated with major adverse cardiovascular events (MACE), such as heart failure, myocardial and cerebral ischemia, may contribute to increased awareness, better management strategies, and potential reduction in morbidity and mortality.

**Inclusion criteria:** This review will include articles describing electrocardiographic changes and the occurrence of MACE in adults with autoimmune rheumatic diseases, except type 1 diabetes.

**Methods:** The protocol will follow the JBI methodology, and the writing will follow the PRISMA-ScR guidelines. The search strategy will be carried out in the PubMed, BVS, CENTRAL, Embase, Periódicos CAPES and Google Scholar databases, including publications from the last 10 years without language restrictions. The Covidence platform will be used in the selection, identification of duplicates, two-stage review and data extraction, conducted by independent reviewers. The data will be analyzed according to the question and objectives of the scoping review, and the results will be presented in tables, charts, and a narrative summary.

**Final considerations:** It is expected that the proposed scoping review protocol will present the general state of scientific evidence and identify gaps to be clarified on the topic.

## Introduction

Autoimmune rheumatic diseases (SARD) represent a heterogeneous group of immune-mediated inflammatory disorders mainly targeting the musculoskeletal system (Moutsopoulos, 2021). Systemic lupus erythematosus, rheumatoid arthritis, systemic sclerosis, psoriatic arthritis, and ankylosing spondylitis are the most common (Makavos et al., 2020) among more than one hundred known autoimmune diseases, that together accounts for a worldwide prevalence between 5-9% (Conrad et al., 2022). Women aged 25 to 40 are the most affected, although SARD can occur in both sexes and in different age groups (Mehta et al., 2023).

Extra-articular involvement in SARD manifests in different tissues and organs, leading to complications that affect quality of life and increases mortality, highlighting the need to manage the consequences of these diseases during treatment (Wu et al., 2022). Cardiovascular involvement in SARD is widely documented with autoimmunity per se recognized as a risk factor, and not just a specific related condition (Conrad et al., 2022), ranging from 1.4 to 3.6 compared to the general population. This risk is equivalent to a 20 mmHg increase in systolic blood pressure (1.26), a 5 kg/m^2^ increase in body mass index (1.29) and the diagnosis of type 2 diabetes (1.62). Despite increasing efforts to develop calculators accounting for the impact of SARD on the cardiovascular system, there is still an underestimation of the risk in these patients, as well as deficiencies in prevention and management strategies, creating a significant gap in comprehensive care for individuals with SARD (Argnani et al., 2021).

The cardiovascular system damage may occur in almost all structures, leading to conditions such as myocarditis, pericarditis, valvular heart disease, early atherosclerosis, heart failure and electrical conduction system disturbance (Prasad et al., 2015). Such changes result from complex mechanisms, triggered by the interaction of genetic and environmental factors (Liu & Perl, 2019). Chronic inflammation plays a central role in this pathophysiological pathway, promoting cardiomyocyte necrosis, structural and electrical remodeling of the myocardium in addition to autonomic dysfunction due to sympathetic hyperactivity and reduced parasympathetic tone (Gawałko et al., 2020).

Cardiac arrhythmias are frequently observed in patients with SARD, including Sinus node dysfunction, atrioventricular block, Conduction tissue disease, atrial and ventricular arrhythmias, which contribute to increased cardiovascular morbidity and mortality (Prasad et al., 2015; Gawałko et al., 2020). Among the main electrocardiographic changes found in the literature, corrected QT interval (QTc) prolongation and increase in QT dispersion (QTd) are associated with a higher risk of ventricular arrhythmias and sudden death. Changes in the P wave, such as increased duration and dispersion, have also been reported, suggesting a predisposition to atrial fibrillation (Prasad et al., 2015). In addition, QRS enlargement, fragmented QRS, and bundle branch blocks are also observed (Gawałko et al., 2020; Argnani et al., 2021). The analysis of the electrocardiogram (ECG), a low-cost and widely available method, is a valuable tool in detecting electrocardiographic changes associated with major adverse cardiovascular outcomes (MACE) in this high-risk population (Conrad et al., 2022; Safdar et al., 2024). However, papers on the subject mostly investigate the cardiovascular effects of just one autoimmune disease at a time, which, together with the small sample size involved, makes it difficult to properly summarize their results (Conrad et al., 2022). This protocol aims to deepen knowledge by aggregating data of a wide range of SARDs, as well as different electrocardiographic abnormalities, to map the main disturbances related to MACE.

A preliminary search was carried out in MEDLINE, Cochrane Database of Systematic Reviews and JBI Evidence Synthesis, and no current or ongoing systematic or scoping reviews on the topic were identified. The research question in the PCC format was developed for this protocol using as a reference the data from Conrad et al. (2022), published in 2022, on 19 autoimmune pathologies and 12 cardiovascular diseases, extracted from a population-based study in England, together with the ACR/EULAR definitions and the main electrocardiographic findings described by Gawałko et al. (2020). It is expected that the scoping review to be generated through this protocol will present the general state of the scientific evidence and identify the gaps to be clarified on the topic.

### Objectives

To present the evidence available in the literature on the presence of electrocardiographic abnormalities and their association with cardiovascular outcomes in patients with autoimmune rheumatic diseases.

### Specific objectives

- Identify the most common types of electrocardiographic abnormalities described in this population.
- Assess the frequency of these abnormalities across different autoimmune rheumatic diseases.
- Investigate the association between electrocardiographic abnormalities and adverse cardiovascular outcomes.

## Methods

The scoping review protocol will be conducted in accordance with the methodology proposed by the Joanna Briggs Institute (JBI) for scoping reviews and will follow the Preferred Reporting Items for Systematic Reviews and Meta-Analyses extension for Scoping Reviews (PRISMA-ScR) guidelines for writing. Since this is a study of publicly accessible secondary data of the scoping review type, assessment by the Research Ethics Committee is not required.

### Research question

The research question was developed using the mnemonic PCC (Population, Concept, and Context), as follows: a) Population (P): adults aged 18 years or older, of both sexes. Studies that included children or individuals diagnosed with diabetes mellitus were excluded; b) Concept (C): systemic autoimmune rheumatic diseases; Context (C): electrocardiographic alterations. The outcomes of interest included major adverse cardiovascular events (MACE), death, stroke, heart failure, and death from cardiovascular causes. Randomized clinical trials (RCTs), observational cohort studies, review studies, and case reports and series were included.

### Eligibility criteria

Eligibility criteria will be established based on the elements of the PCC model (Table 1):

**Table 1:** Study eligibility

|  |  |
| --- | --- |
| **Population** | Individuals with rheumatic diseases, autoimmune diseases, systemic lupus erythematosus, rheumatoid arthritis, ankylosing spondylitis, psoriatic arthritis, psoriasis, polymyalgia rheumatica, granulomatosis with polyangiitis, Takayasu's arteritis. |
| **Concept** | Electrocardiography, electrocardiogram, electrical abnormalities, conduction system, atrial fibrillation, ventricular tachycardia, cardiac conduction system, rhythm, atrioventricular block, extrasystole, QT interval, heart rate variability. |
| **Context** | Major cardiovascular events, major adverse cardiovascular events, heart failure, stroke, sudden cardiac death, myocardial infarction, mortality. |
| **Study/source of evidence selection** | Clinical trials, systematic reviews, cohort studies, series and case reports |

Studies with pediatric populations or with non-autoimmune diseases will be excluded. Clinical trials, systematic reviews, cohort studies, series and case reports, in Portuguese or English, published in a ten-year interval, will be included.

### Information sources and search strategy

Searches will be conducted in the following electronic databases: PubMed, Biblioteca Virtual em Saúde (BVS) – which includes the LILACS, BIREME and MEDLINE databases –, the Cochrane Central Register of Controlled Trials (CENTRAL) repository, the Embase database (Elsevier), accessed through the CAPES Journal Portal, as well as the CAPES portal itself. To broaden the scope of the review, gray literature sources, such as Google Scholar, will also be explored. Study screening and data extraction will be performed through the COVIDENCE platform.

Key search words included but were not limited to the following:

**Table:**
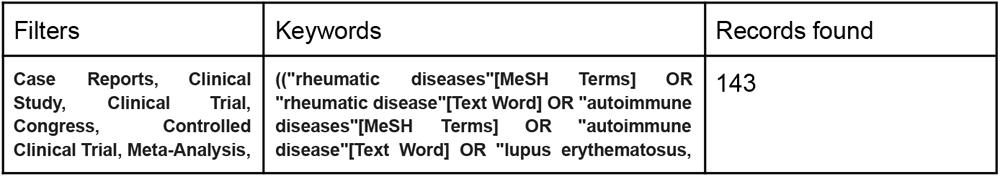
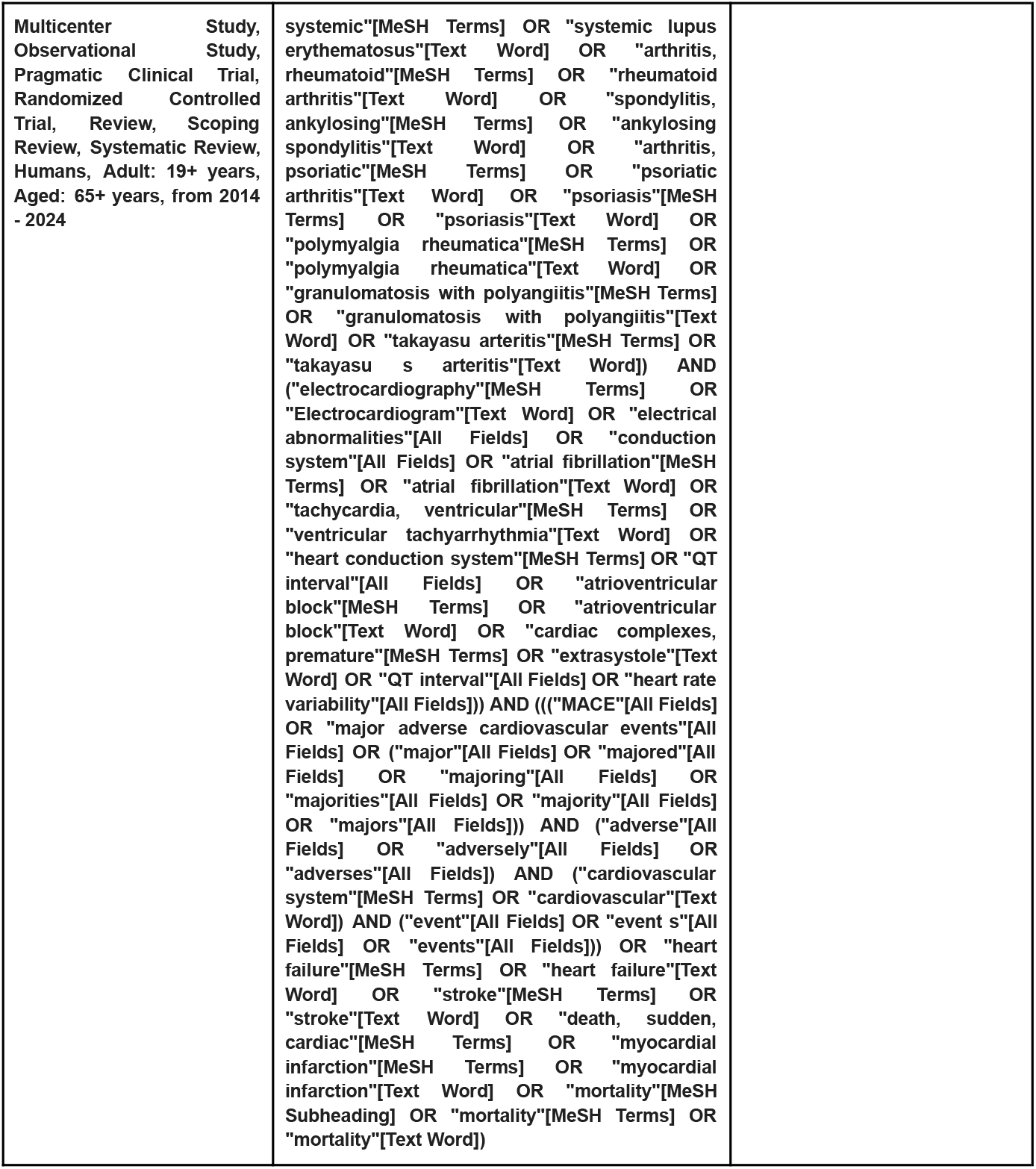
Search strategy - Medline via PubMed

Additional relevant studies, published through the protocol development process, will also be considered by this research group and added to the evidence tables when appropriate.

### Data Selection and Extraction

The selection of studies will be carried out with the help of Covidence software (Veritas Health Innovation, Melbourne, Australia), which will allow the import of references, as well as the identification of duplicates and the management of the review stage. This review will be carried out by removing duplicate records and screening the titles and abstracts by eight independent reviewers, allowing the categorization of studies as included or excluded, using the same Covidence software.

Once this stage is complete, the articles considered eligible will undergo a full reading to assess compliance with the inclusion criteria that were previously defined. The extracted data will then be analyzed according to the objectives of the review, and the results will be presented at the end of the scoping review.

### Outcomes

Variables related to the type of study presented, target population, language and year of publication will be collected. From the selected articles, the outcomes of death from all causes, cardiovascular death, arrhythmias, stroke and heart failure of each study will be analyzed, in addition to the electrocardiographic changes presented.

### Data synthesis

The data will be examined based on the research objectives, highlighting the variables investigated and the methods chosen for the studies. The analysis will be conducted in both quantitative and qualitative ways. The information collected will be presented in a flow diagram, and the most relevant results will be addressed in the discussion to clarify the central question of the study and meet the established objectives.

## Discussion

This scoping review protocol will allow for a comprehensive review of the published literature on Cardiovascular outcomes and ECG abnormalities in systemic autoimmune rheumatic diseases, which has implications for research and policy recommendations.

A wide, multiple language search approach, including the main traditional and key databases along with Google Scholar, covering an extensive number of publications is a strength of this protocol. Despite that, articles not indexed in these databases may be missed. When conducting a literature review on rare diseases, as SARDs, other sources of gray literature, such as reports from relevant organizations on rare diseases, would still be of great value.

A critical appraisal to assess the quality and risk of bias in included studies was not planned, as the main goal of the present review is to explore a broad research area covering multiple study designs.

## Conclusion

The proposed scoping review protocol has the potential to provide a significant contribution to the existing body of evidence regarding electrocardiographic abnormalities and cardiovascular outcomes in autoimmune rheumatic diseases. Given the relevance of the findings to multiple audiences, including healthcare professionals, researchers and academics, the results will be widely disseminated.

“The authors received no specific funding for this work.”

## Data Availability

All data produced are available online at OSF: DOI 10.17605/OSF.IO/G6QDT

